# Human mobility and sewage data correlate with COVID-19 epidemic evolution in the metropolitan area of Bologna

**DOI:** 10.1101/2025.03.27.25324700

**Authors:** Francesco Durazzi, Enrico Lunedei, Giulio Colombini, Giulia Gatti, Vittorio Sambri, Alessandra de Cesare, Cecilia Crippa, Frédérique Pasquali, Bologna MODELS4COVID Study Group, Gastone Castellani, Daniel Remondini, Armando Bazzani

## Abstract

The COVID-19 pandemic has significantly impacted human society at many levels, from public health to economics and transports, highlighting the need of approaches integrating all available information to better understand and model similar phenomena, also in order to develop early detection and responses. In this paper we show the result of an analysis of COVID-19 pandemic in the metropolitan area of Bologna, Italy, integrating an epidemiological mathematical model, SARS-CoV-2 virus quantification in wastewater, clinical hospitalization, vaccination campaign, virus geno-typization and human mobility data in the period 2020-2022. We were able to follow the evolution of epidemic, observing the effect of vaccination and other factors that produced significant changes in hospitalizations. Moreover, by considering a mathematical model of COVID-19 epidemics spread, with parameters selected partly from literature and partly adapted to the local situation on a weekly basis, we identified a strict relation between human mobility at mesoscopic level and a sociability rate (related to model reproduction number) revealing the mutual impact of health issues on human activity and viceversa.

## 1 Introduction

The COVID-19 epidemic is the first large pandemic globally monitored with an enormous amount of collected information and data from different sources. Since SARS-CoV-2 was detected in December 2019, much effort has been put into disease modeling, to inform the pandemic response. Given the extreme speed of spread on a global scale, attempts at forecasting have been accompanied by a compelling need for *nowcasting*, trying to understand and quantify the extent of the contagions on short time scales. With enough data, predictive models proved to be quite effective, showing in the U.S. an average hold of 22 weeks, barring the emergence of unforeseen new mutations[1]. Human mobility data has often been considered an informative data source for epidemiological models[2, 3, 4, 5], though in the context of COVID-19 epidemic it has been observed that the relative explanation added by mobility time series to predict future epidemic trends was relatively small[6] and changed over time[7]. Epidemiological modeling also helped estimate the effect of interventions aimed at reducing the number of transmissions. In Italy, one of the countries mostly hit by COVID-19 in Europe, a compartmental model informed by mobility data estimated that restrictions reduced transmission from 42% to 49%[8]. Traditional surveillance based on clinical observations has been supported by real-time genomics aimed to identify the virus and follow the emergence of new variants. Intense sequencing efforts have made it possible to reach more than 100,000 complete genomes on GISAID as early as the end of 2020[9], enabling real-time transmission monitoring and informing public health decision-making [10, 11, 12]. The pressing demand for data and information led to consideration of any data source that could help shedding light on the pandemic, even involving online social networks[13] and app-based contact tracing[14]. In the search for an optimal trade-off between health and privacy, a popular practice has been performing measurements based on environmental samples. Next-generation sequencing applied to wastewater samples operated in complementarity to disease-based surveillance, taking advantage of the fact that 40% of infected people shed the virus in their feces[15]. Through an appropriate sewage surveillance system, it is possible to monitor the presence of the virus in the city in nearly real-time, overcoming the bias attached to the number of positive tests, which depends on test availability and indications[16, 17, 18]. In Milan, Italy, the correlation between wastewater SARS-CoV-2 load and the number of hospitalizations has been declining over time in correspondence with increasing vaccination coverage, thus showing the effectiveness of vaccinations in increasing population immunity[19].

In our work, we integrated wastewater epidemiology with a mathematical model of infections and hospitalizations and with several metropolitan-scale data sources, to explore the capacity of a multi-modal approach to cope with a developing epidemic. Over an observation period ranging from 2020 to late 2022, for the metropolitan area of Bologna we integrated sewage sequencing, pathogen genotyping and human mobility data, and then compared them with time series of COVID-19 tests, vaccinations, and hospitalizations. In our results 1) we derive a quantitative relationship between wastewater viral load and clinical observations related to positive cases and hospitalizations on a long time scale, and observe how this relationship varies over time in correspondence to macroscopic factors such as vaccination coverage and significant virus mutations; 2) using a predictive mathematical model informed by clinical observations, and parameters partially inferred from literature, we show how tuning the model to real data underscores a correlation between the social contact parameter in the model and measures of urban mobility at high time resolution, which could then be used as a proxy for parameters to be fed into epidemiological models.

## 2 Methods

### 2.1 Clinical COVID-19 data

The number of new COVID-19 cases, full vaccinations and hospitalizations were provided by the Bologna health agency ASL, and refer to the Bologna metropolitan area (about 900,000 inhabitants)[20]. Data is used with a daily resolution to inform a compartmental model for COVID-19 spread (described below) and averaged to a biweekly resolution for the comparison with urban sewage data. The positive test ratio is defined as the ratio between weekly new cases and tests, and was obtained by rescaling the number of tests in the Emilia Romagna region to the population size of Bologna.

The time series of tests in Emilia Romagna region was obtained from the data repository of Civil Protection (Protezione Civile, https://github.com/pcm-dpc/COVID-19). Information about the timeline of infection control measures and lockdowns in Italy can be found at https://lab24.ilsole24ore.com/storia-coronavirus/.

### 2.2 Virus genomic data

To gather information about the emergence of SARS-CoV-2 variants in the city, we considered a dataset of 1,060 SARS-CoV-2 isolates sampled from human patients by the Operative Unit of Microbiology of The Great Romagna Area Hub Laboratory, Pievesestina (FC, Italy). All SARS-CoV-2 nasopharyngeal swabs were tested through the real-time Polymerase Chain Reaction (PCR) Allplex SARS-CoV-2 Extraction-Free assay (Seegene Inc., Seoul, Republic of Korea) targeting E, RdRP/S and N genes. Among positive leftover samples, random nasopharyngeal swabs were selected to be sequenced from March 2021 to May 2022 and during the daily routine. Collected samples were anonymized according to laboratory’s procedures to adhere to the regulations issued by the Local Ethical Board (AVR-PPC P09, rev.2; based on Burnett et al., 2007). The genetic material was extracted through the automated system Maelstrom 9600 (TANBead—Taiwan Advanced Nanotech Inc., Taiwan). Then, the library was prepared using the CleanPlex SARS-CoV-2 Flex Research and Surveillance NGS Panel assay (Paragon Genomics, Inc., Hayward, CA, USA) and sequenced on the MiSeq platform (Illumina, San Diego, CA, USA). Forward and reverse FastQ files were uploaded and analyzed on SOPHiA DDM software (SOPHiA GeneticsTM, Lausanne, Switzerland), aligned on the Wuhan reference genome (NCBI Accession number: NC 045512.2) to generate a consensus sequence. We extracted 230 unique Spike protein sequences from the available genomes, and aligned them against the main SARS-CoV-2 lineages (up to Omicron BA.2) for this study. We used the date of collection of the first BA.1 sequence observed in the dataset as a reference about the emergence of the Omicron variant in the region, which determined a significant change in the pandemic evolution worldwide both genetically and clinically[21]. The laboratory’s coverage area (Romagna) does not include the city of Bologna itself, but it covers an area ranging between 50km and 130km from the center of the city of Bologna and is located within the same administrative region of which Bologna is the capital (Emilia-Romagna), sharing its health administration. Given the close proximity, daily commuter traffic, and the rapid onset of emerging variants, it is reasonable to assume that the arrival time of the various lineages does not differ much between the two areas.

### 2.3 Urban mobility data

The mobility data used in this work mainly originates from the “Open Data - Bologna” project of the Bologna municipality[22] (https://opendata.comune.bologna.it/explore/dataset/rilevazione-autoveicoli-tramite-spire-anno-2021/), which is a large open access repository containing several datasets and, in particular, the hourly traffic flows data from 292 magnetic coils distributed in the roads of Bologna metropolitan area. The available data covered the full span of years between 2020 and 2022. Our hypothesis is that the traffic flow time series can be used as a proxy of the social activity in the city, which is one of the factors contributing to the COVID-19 epidemic spread, and thus that the changes in the traffic flows (easily measurable) can be correlated to changes in the social activity (less easily measurable). We considered the daily traffic flow time series in Bologna smoothed by a 7-day moving average, in order to reduce the traffic fluctuations during the weekly cycle of mobility. Finally, the total traffic flow was rescaled to the value at the onset of COVID-19 pandemic, and this rescaled value constituted our mobility index *m*.

### 2.4 Viral load of SARS-CoV-2 in urban sewage

Untreated sewage samples were collected approximately twice per month since November 2020, from the urban sewage treatment plant covering the whole city of Bologna. Up to November 2022, 38 samples were collected in total. Upon arrival to the lab, fresh, not frozen, samples were immediately submitted to filtering and subsequent RNA extraction. In particular, two aliquots of 15 ml each of the sewage supernatant were placed in Amicon Ultra 15 30 kD tubes (Millipore) and centrifuged for 40 min 4000 xg at 4 ^*°*^C. The filters were then rinsed with 200*µ*l of Phosphate Buffer Saline (PBS) solution, which was then submitted to RNA extraction using the QIA AMP viral RNA mini kit (Qiagen, Milan, Italy). RNA samples were tested by reverse transcription real-time PCR targeting the E gene of SARS-CoV-2 virus following previously reported protocol (Corman et al., 2020) on Applied 7500Fast Real-Time PCR systems (Termofisher, Milan, Italy). In particular, reverse transcription was performed with TaqMan Fast Virus 1-step Master Mix (Thermofisher, Milan Italy) in a total volume of 20*µ*l. A standard curve was built on serial dilutions of SARS-CoV-2 RNA of the reference sample. Based on Ct values, the equation of the standard curve *y* = *−* 4, 3773*x* + 55, 375 (*R*2 = 0, 9683) was used to calculate the initial viral concentration (log_10_RNA copies/*µ*l) in sewage samples. This concentration value over time is what we refer to as *viral load in urban sewage*. When missing, the time series was linearly interpolated at biweekly resolution, to be compared with clinical data and model output.

### 2.5 Compartmental model for epidemic spread

To model the COVID-19 epidemic spreading in the Bologna metropolitan area, we used a compartmental SEUIR model (Susceptible - Exposed - Unreported infected - Isolated infected - Recovered) based on delay differential equations[23, 24, 25, 26], that extends the traditional SEIR model (Susceptible-Exposed-Infected-Recovered[27]) with a compartment devoted to the unreported infected population, which plays a key role in the SARS-CoV-2 epidemic spreading. We considered a SEUIR model with delay due to the significant gap between infection and appearance of symptoms, allowing the identification of infected people and the mandatory self-isolation, characteristic of COVID-19 infection, as in [28]. Specifically, for any time *t ≥* 0, the population is subdivided as *N* (*t*) = *P* (*t*) + *I*(*t*) where *N* is the total population in the area, which is assumed as constant except for the number of dead individuals. The total population is divided into “active population” (*P*) who contributes to social activities and disease spreading, and “infected individuals put in isolation” (*I*), either at home or hospitalized, which contribution to the epidemic spread we approximated to negligible. The active population *P* is further divided into four compartments (*P* (*t*) = *S*(*t*) + *E*(*t*) + *U* (*t*) + *R*(*t*)), where *S* is the number of *Susceptible* individuals, *E* is the number of *Exposed* individuals (i.e., people that are positive to the virus, but are still not contagious), *U* is the number of *Unreported* infected individuals (i.e., infected individuals without evident symptoms of the disease, who are contagious and not reported) and *R* is the number of *Recovered* individuals that are immune to contagion. The *R* compartment contains both the healed individuals and the vaccinated individuals. Although both *R* and *I* individuals do not actively contribute to disease spread (being healed or isolated) they do have a different role, since Recovered individuals are part of the active population *P* (thus diluting the fraction of Unreported infected), while Isolated infected are not. We did not consider the age structure of the population in the model since we did not have sufficient available data.

The evolution of the epidemic is simulated as follows (see the diagram in Figure 1):

**Figure 1:**
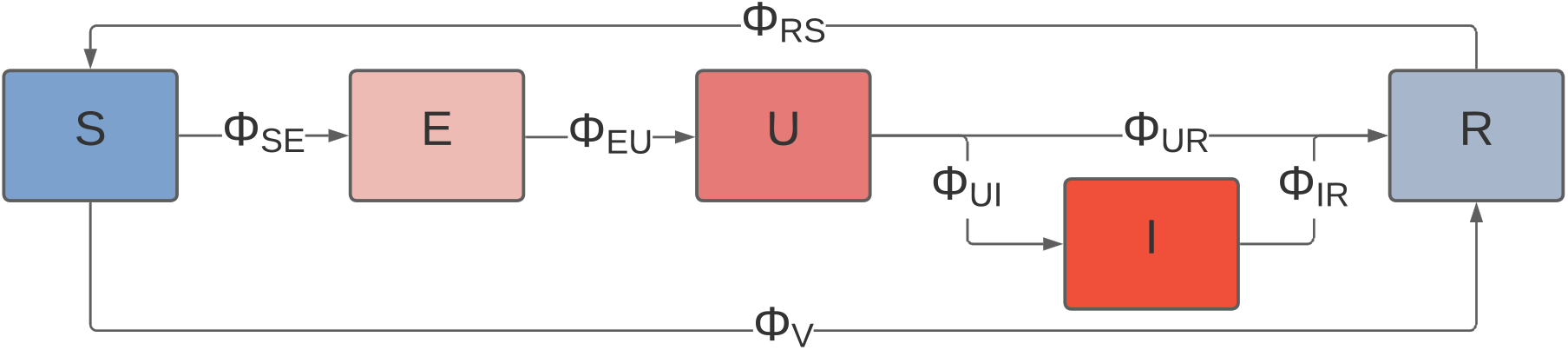
Compartmental model scheme, describing equations in (1).

1. a *Susceptible* individual may become *Exposed* after meeting an *Unreported* infected individual. The flow of new infections is controlled by the factor *β*, which is the rate at which potentially infectious contacts occur[29]. *β* is modulated by two multiplicative parameters, namely *τ* (*relative infectivity w*.*r*.*t. the initial SARS-CoV-2 variant*), accounting for virus evolution over time, and *s* (*relative sociability*), which describes the variation of the average number of social contacts with respect to its initial value contained in the *β* parameter (details in Supplementary Text);
2. after a time interval *T*_*E*_, an *Exposed* individual becomes an *Unreported* infected one;
3. an *Unreported* infected individual can infect other people and they can, with a probability *α*, develop the symptomatic disease and be isolated or, with a probability (1 *− α*), recover and be transferred to the *Recovered* compartment after a time interval *T*_*U*_;
4. reported infected individuals are *Isolated* at home or within a hospital ward and, after some time, are moved to the *Recovered* compartment (or they die). For the sake of simplicity in the paper we considered a time *T*_*I*_ corresponding to the quarantine period;
5. after a time interval *T*_*R*_, *Recovered* individuals return to the *Susceptible* compartment, as the natural or vaccine-induced immunization is lost.

The average flux between Susceptible and Exposed compartments is

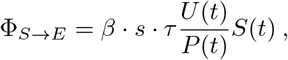

where one assumes the probability of meeting an infected unreported individual *U* proportional to their prevalence in the population, since urban mobility diffuses the individuals in the whole metropolitan area (homogeneous mixing). Using two distinct multiplicative factors (*s* and *τ*), we disentangle infectivity and sociability, and explicitly consider the role of social activities that change during the pandemic due both to social restriction measures introduced by the government and people behavior.

The compartmental model is defined by the delay differential equations (see model scheme in Fig.1)

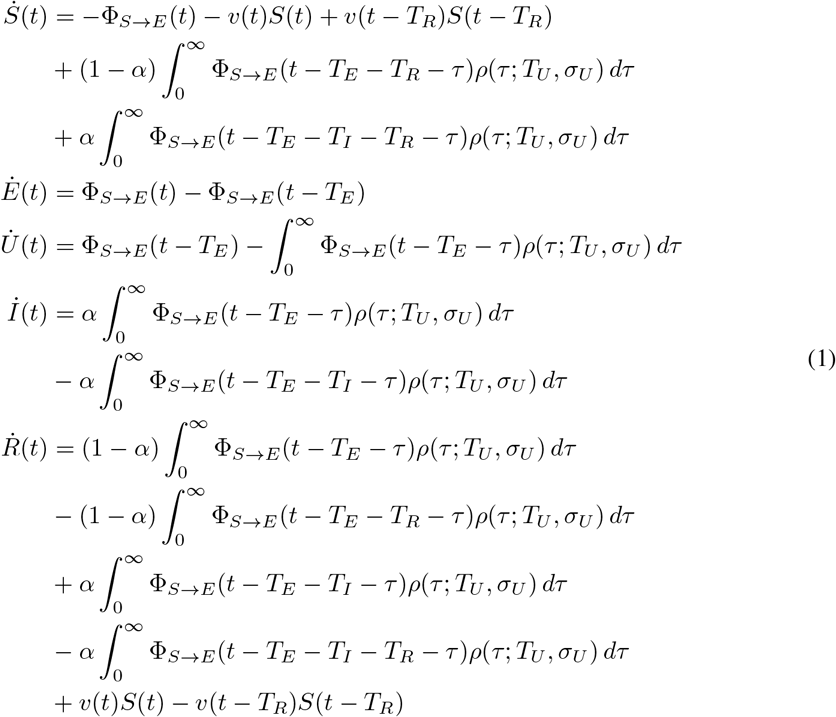

where *ρ*(*τ*; *T, σ*) is the Gamma-distribution function with average value *T* and variance *σ*^2^

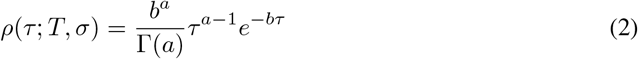

where *a* = (*T/σ*)^2^ and *b* = *T/σ*^2^ and Γ(*a*) is the Gamma function. The permanence time values for the compartments, reported in Table 1, have been taken from literature[30]. Only the *U* delay has been considered in its distributed form, while all other delays have been treated as discrete, their value corresponding to the mean permanence time observed in literature. The fraction of reported infected *α* was set to *α* = 0.14 according to literature[28].

**Table 1:**
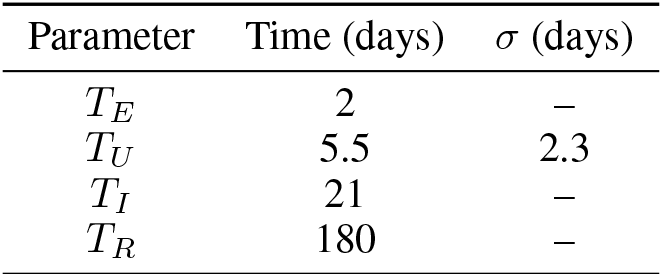
Permanence times of each compartment in the model. The standard deviation *σ*_*U*_ is reported only for the compartment where it is defined.

At the initial condition of the model *s* and *τ* have been set to 1, and the value of *β* parameter has been estimated with a linear fit of the logarithm of the infection data time series of the first 10 days (24/02/2020 - 04/03/2020, assuming an exponential growth as typical in epidemiologic models). For compartmental models such as (1), the relationship between *β* and the basic reproduction number *R*_0_ can be calculated as *R*_0_ = *β · T*_*U*_ [23]. The estimated value *β* = 0.833 *±* 0.003 from our data produces an *R*_0_ compatible to wild-type values found in literature (*R*_0_ = 4.6 *±* 1.9, with uncertainty mainly due to *T*_*U*_ but with *β* estimated on very few data points, to be compared to a 95% confidence interval of [2.4 *−* 3.4][31]). The relative infectivity *τ* has been modified three times, due to the increased infectiousness of emerging SARS-CoV-2 variants with respect to the wild-type: *τ* = 1.56 since the 20^th^ of October 2020 (estimated arrival time for the Alpha variant), *τ* = 2.50 since the 1^st^ of April 2021 (Delta variant), and by *τ* = 12.5 since the 1^st^ of December 2021 (Omicron variant). These values have been set accordingly to the news released by the Italian health institute (ISS, https://www.iss.it/cov19-cosa-fa-iss-varianti), and are not inferred through the model.

The relative sociability parameter *s* was varied on an approximately weekly basis during two years from the beginning of COVID-19 epidemic, in order to minimize the difference between predicted and observed new cases of confirmed infections (i.e., considering the number of individuals moving from the Unreported to the Isolated compartment), and *s* is the only free parameter of the model. Since the best fit value was obtained by varying the *s* parameter in steps of 0.01, this value also represents the uncertainty associated to the *s* parameter (see also Supplementary Text). Finally, the effect of the vaccination campaign was modeled through the term Φ_*V*_ (*t*) = *v*(*t*)*S*(*t*), with *v* the parameter quantifying the effect of vaccination in reducing the susceptible population *S*. The vaccination parameter *v*, which lowers the number of susceptible individuals because of immunization, was set to 0 up to the 1^st^ of January 2021 and its value was afterwards updated to the vaccination rates provided by the local health unit on a weekly basis. In the model we hypothesize that vaccine efficacy is 100%, and we show in Supplementary Text the results of the analysis with a reduced (75%) vaccine efficacy.

### 2.6 Modeling the relationship between clinical and sewage viral load data

To characterize the relationship between the experimental observations of SARS-CoV-2 viral load in sewage, positive test ratio and hospitalizations over the three phases, we designed a linear model with the following equations:

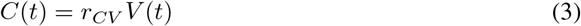

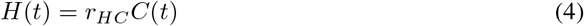

where *V*, *C* and *H* are the sewage viral load, the COVID-19 positive tests ratio, and the number of hospitalizations respectively, sampled on a biweekly basis. The choice of a linear model is motivated by the significant linear correlation between the time series (see Results section and Supplementary Figure S2). The model assumes no time lag between the time series; this assumption is motivated by the fact that we estimated the time lags providing maximum linear correlation both between 1) sewage viral load and COVID-19 positive tests ratio, and 2) between hospitalizations and COVID-19 positive tests ratio, and they resulted equal to 0 weeks. We used Monte Carlo Markov Chains (MCMC) to parametrize the model through the Python’s package pymc3, which also allows to inspect the a-posteriori distribution of the parameters. The Monte Carlo sampling consisted of 4 chains of 1000 NUTS steps each with acceptance probability of the proposed samples set to 0.85, and a-priori parameters value set to the average ratio of the observed time series: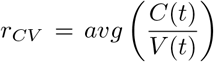 and 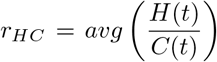. We then characterized the variations of the estimated ratios over time, exploring the relationship with factors such as vaccination rate in the population and the emergence of new SARS-CoV-2 variants.

### 2.7 Modeling the relationship between sociability and mobility index

We compared the sociability index *s* inferred through the compartmental model with the mobility index *m* obtained from urban mobility data. First, we rescaled both indices with respect to their maxima occurring before the pandemics (as shown in the first white region in Fig. 4a). Second, the indices were aligned through a translation of the values on the *y* axis (shift) guided by a) an increasing discrepancy between *m* and *s* indexes, and b) the close occurrence of specific events like re-openings, or the emergence of virus variants with different properties (e.g. the Delta variant) that could have altered the relationship between human mobility and sociability. The relevance of this breakpoints was also validated *a posteriori* through a statistical change-point analysis (see Supplementary Text). We then evaluated in each of these time periods the correlation between the two time series and computed the mean squared error *MSE* in predicting the sociability parameter *s* using the mobility index *m*.

## 3 Results

### 3.1 Viral load in urban sewage

We compared the number of COVID-19 infections and hospitalizations with the viral load measured in urban wastewater, to help us characterize the major factors during the epidemic that changed the relationship between the effective presence of the virus in the population and the available clinical observables. In Figure 2, we show the SARS-CoV-2 viral load in urban sewage over time, together with the positive tests ratio, and the number of daily new cases and hospitalizations.

**Figure 2:**
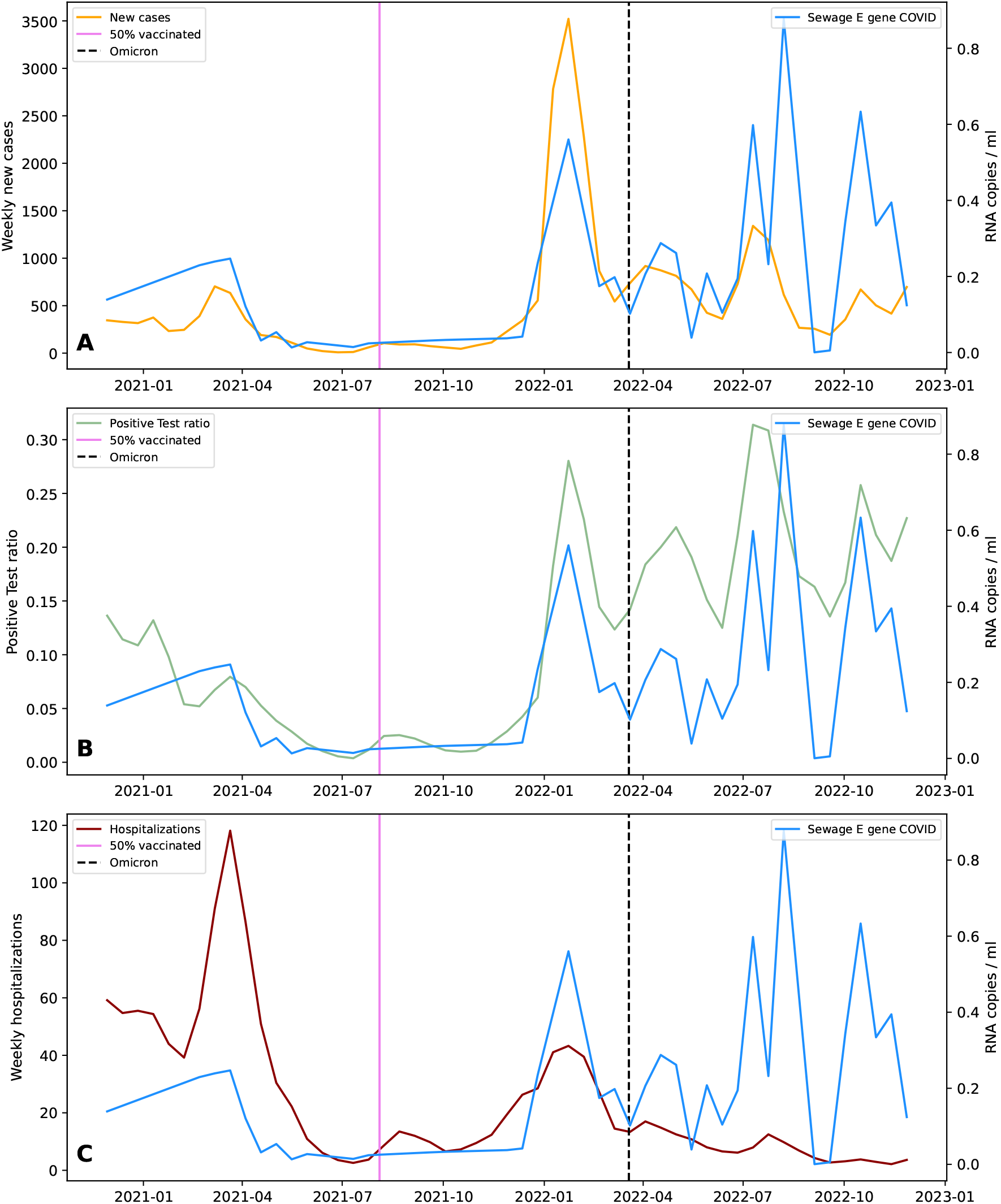
SARS-CoV-2 viral load in urban sewage (blue), plotted with a) number of daily new COVID-19 cases (orange), b) average weekly positive tests ratio (green), c) number of daily COVID-19 hospitalizations (red). The pink vertical line represents the reach of 50% vaccination coverage in the population. The vertical dashed black line represents the date in which the first Omicron BA.1 sequence was retrieved from a patient in our dataset.

The agreement between the sewage viral load and the number of cases is good throughout all the observed period (Pearson’s correlation coefficient r=0.57, p-value *<* 10^*−*6^), in particular considering the position of the main peaks of COVID-19 infections on March 2021, January 2022, April 2022 and July 2022 (Fig. 2a). The ratio of positive tests (Fig. 2b) correlates better with the sewage viral load (r=0.73, p-value *<* 10^*−*6^). The biggest difference between the number of new cases and the positive tests ratio occurs during the peaks of July and October 2022, in which the positive tests ratio is as high as in the peak of February 2022, while the number of new cases is lower than in February even with a similar dynamics. The number of COVID-19 hospitalizations (Fig. 2c) decreased peak after peak, in accordance with the increase in the percentage of vaccinated people first, and the arrival of Omicron variant later, while the peak amplitude of sewage viral load remained constant for the whole 2022. In facts, its correlation with cases is not significant (r=0.19, p-value=0.17) reflecting the variation of the relation over time.

The vaccination campaign and the appearance of the Omicron lineage can be used to identify three different phases: phase A, up to the 50% of vaccinated individuals on 1^st^ August 2021; phase B, from the end of phase A up to the arrival of Omicron BA.1 lineage on 19^th^ March 2022; phase C, after the Omicron lineage became prevalent since March 2022. We thus fitted the MCMC linear model in (4) to the three phases separately (see Supplementary Table S1 for values, and Supplementary Figure S3 for details on the convergence of the method). In Figure 3, we show the estimated parameter values over the three time phases.

**Figure 3:**
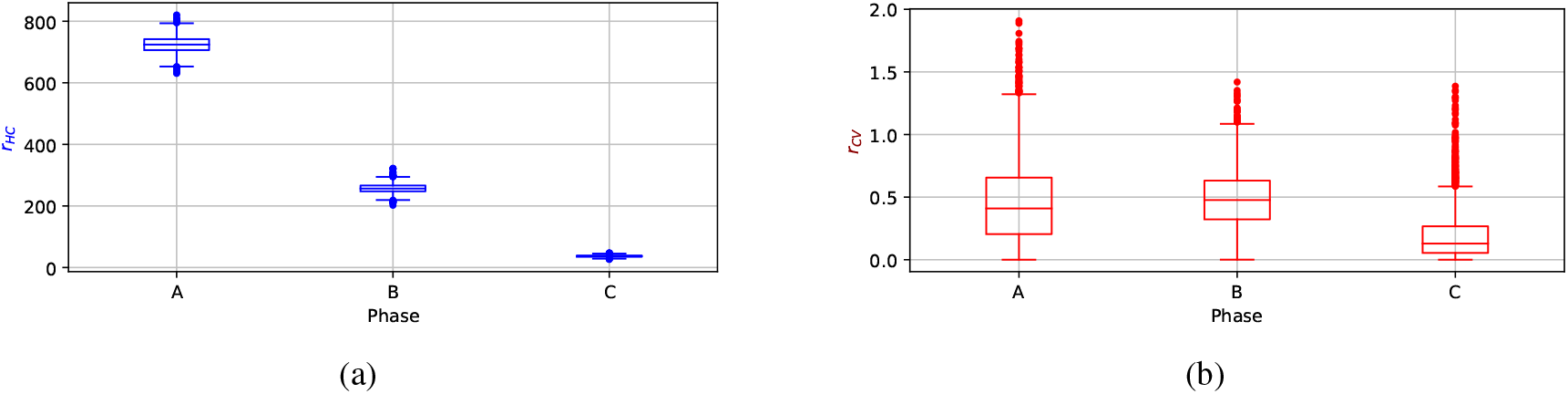
Boxplot of fitted parameter distributions over the time phases, for the MCMC model. a) *r*_*HC*_ (blue) is the ratio between COVID-19 hospitalizations and positive tests ratio. b) *r*_*CS*_ (red) is the ratio between COVID-19 positive tests ratio and SARS-CoV-2 sewage viral load.

In the B phase, the ratio between the number of hospitalizations and the percent of positive tests (*r*_*HC*_) is less than half its value during phase A, while the ratio between the percentage of positive tests and sewage viral load (*r*_*CS*_) remains constant, possibly because in phase B more than half of the population was vaccinated, thus reducing hospitalization risk even if viral load did not decrease. In phase C, *r*_*HC*_ is drastically reduced (around 1*/*20 than in phase A), and we hypothesize that it is due to the effect of the vaccination campaign added to the lower severity of Omicron lineage, which became prevalent since then. The correlation between the percentage of positive cases and sewage viral load is significantly reduced (r=0.66, p-value=0.055 in phase C as compared to r=0.80, p-value *<* 10^*−*6^ in phases A and B together), and also *r*_*CS*_ decreases (Mann–Whitney U test p-value *<* 10^*−*6^), showing a decoupling between clinical tracing and the presence of SARS-CoV-2 in sewage (see also Supplementary Figure S2 for scatter plots in the three phases).

### 3.2 Sociability and traffic data

Regarding the compartmental model in (1), we fitted the time series of the *I* compartment to the observed number of newly infected cases by tuning only the value of *s* parameter. This procedure was applied in the Metropolitan area of Bologna during the COVID-19 pandemic on a weekly basis, to support a health crisis unit assisting the local hospital agency. In Supplementary Figure S1 we show how the model (1) was able to reproduce the observed new reported infected cases during the first two years of COVID-19 epidemic.

This procedure of continuous adjustment allowed us to obtain a time series of the sociability parameter *s* inferred from epidemiological data, which can be interpreted as a measure of human interactions during the pandemic. In Fig. 4a we compare the sociability parameter *s* and the mobility index *m*, the latter derived independently from the traffic flow time series measurements in Bologna Metropolitan area (see Methods).

With the enactment of the first national lockdown (March 2020, first red-shaded area in Fig. 4a), the traffic flow dropped sharply to the *≃* 20% of the initial pre-pandemic value. Afterwards, several restrictions remained in place, and the observed mobility index, though increasing, remained lower than the pre-pandemic value (about 80% in June 2020). The introduction of curfews and pandemic severity-modulated measures on a regional scale (Autumn 2020) corresponds to the second red-shaded area in Figure 4a and it reduced the mobility index up to 70% of the initial value. Another reduction in mobility happened during March and April 2021; indeed this period corresponds to the application of some restrictive rules for social activities by the Italian government (third red-shaded area). Other major oscillations are observed in correspondence with the summer holidays (August 2020 and 2021) or the end of the year (Christmas holidays), represented as blue-shaded areas in Figure 4a The mobility decrease during holidays can be also the consequence of the population leaving the Bologna area, and for the Christmas holidays it is often preceded by a short increase in mobility in correspondence with Christmas shopping. During the whole observation period, one can also recognize a global trend towards the recovery of the initial pre-pandemic mobility, which is approximately restored starting from December 2022.

At the beginning of the first lockdown (white shaded area in Figure 4a), the mobility index *m* had the same behavior as the sociability parameter *s*. However, when the mobility restrictions were partially removed in May 2020 we observed a systematic discrepancy between them. In fact, while the mobility index increased to *≃* 80% of its initial value, sociability did not recover to the same extent. This mismatch could be removed by shifting the value of *s* by an additive value of 0.58 (see Table 2), as we show in Figure 4b. A possible explanation of this discrepancy between the two indices could be the drastic change in people’s behavior during their social activities, like the obligation to use health masks in many contexts, thereby reducing the infection probability despite the increase in social contacts.

**Table 2:** Breakpoints for the correlation analysis between sociability and mobility indexes. The values in the Shift column were added separately (not cumulatively) to the baseline sociability index.

| Day | Event | Shift |
| --- | --- | --- |
| 18/05/2020 | Activities reopening (bar, restaurants) | 0.58 |
| 17/05/2021 | Delta variant in Emilia Romagna | 0.23 |
| 15/09/2021 | Schools reopening | 0.71 |

The agreement extends for about one year, after which the effect of the vaccination campaign, the arrival and prevalence of the Delta variant, and the beginning of summer changed the epidemic spread dynamics (black vertical line in in Fig. 4b), requiring a smaller shift of the sociability parameter *s* equal to 0.23 to make the two indices match. The smaller discrepancy can be explained by a reduced usage of social distancing and masking, likely due to warmer weather and changes in social behavior in summer.

**Figure 4:**
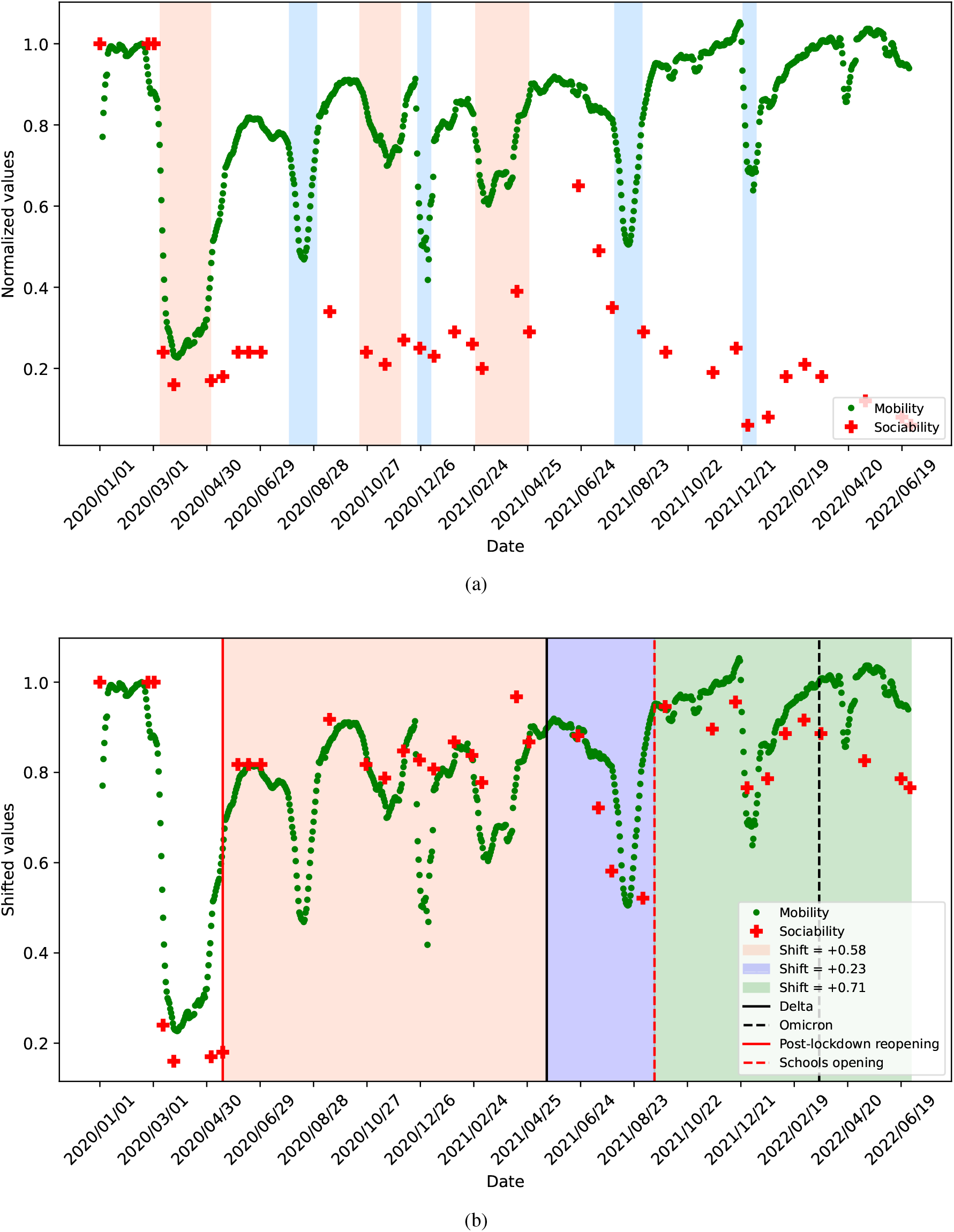
Comparison between the sociability parameter *s* (red crosses) and the mobility index *m* (green dots). a) The red-shaded areas mark 1) the Italian national lockdown during the first epidemic wave (3^rd^ March - 4^th^ May 2020), 2) government-mandated countermeasures during the second pandemic wave, such as the shutdown of commercial activities and the introduction of night curfews and color-coded regions (19^th^ October 2021 - 3^rd^ December 2020), and 3) winter restrictions from 26^th^ February to 26^th^ April 2021. The blue-shaded areas mark winter holidays and the month of August. b) The shifted piece-wise function *s* (red crosses) is plotted against the mobility data (green dots). The shaded regions refer to different shifts applied to the sociability rate (see legend).

After summer 2021, when a resurgence of the epidemic occurred after schools reopening (see Table 2) we shifted the sociability index *s* of a factor equal to 0.71. We hypothesize that this greater difference between *s* and *m* parameters might reflect other factors affecting COVID-19 epidemics. The effect of immunity due to vaccination and the reduced susceptibility of the population to the newly emerging virus variants reduced the amount of infections (represented by parameter *s*) with respect to the actual real number of contacts (represented by the parameter *m*).

With the three introduced shifts (shown in Figure 4b) the two time series resulted significantly correlated (Pearson’s coefficient *r* = 0.78, p-value *<* 10^*−*6^), thus the sociability index *s* could have been estimated from the mobility index *m*, more easy to obtain from on-site measurements of road traffic, with a Root Mean Squared Error *RMSE* = 0.15. For comparison, a null model, in which we performed 1000 random extractions of 3 breakpoints in which to perform the shift, achieved a significantly lower average Pearson’s correlation coefficient *r*_0_ = 0.50 *±* 0.15 and a significantly higher error *RMSE*_0_ = 0.32 *±* 0.07. In the Supplementary Text, we show for comparison the results of a statistical change point detection analysis operated on the difference between the sociability parameter and the mobility index. The analysis highlights 4 break points: the first 3 are close (within a month) to the ones we selected based on significant external events, while the additional one overlaps with the arrival of Omicron lineage in the region, which impacted again the relationship between sociability and mobility. Specifically, the sociability parameter started to decrease since early 2022, and its correlation with the mobility index gradually reduced. As we observed from the comparison with the sewage data, the emergence of the less severe Omicron variant and a high degree of vaccination coverage caused a decoupling of the number of COVID-19 cases from the virus abundance in wastewater. In this last phase, the number of positive tests is so low that it was no longer reliable to feed the compartmental model.

## 4 Discussion

In this study we show how the integration of different data sources can support surveillance of the spread of an infectious pathogen over an urban area. Indeed, the intersection of data offering different perspectives provides greater explanatory power than single-source studies and allows for the identification of different phases in the course of the epidemic. In our case study, we addressed the COVID-19 epidemic with data from the Metropolitan area of Bologna, Italy, over a period from January 2020 to November 2022. Specifically, we monitored the official number of infections and hospitalizations through clinical data, the appearance of different virus variants through genomic sequencing, the actual presence of the virus in the population through urban sewage sequencing, and the social activity of the population through road vehicle mobility.

The first interesting result is the statistically significant correlation between the level of road traffic and the social activity parameter inferred from the epidemiological model, which motivates the possibility of using aggregated urban mobility data as a proxy to inform high-temporal resolution predictive models about social contacts. During an epidemic, new control measures are continually introduced and updated to modify population behavior in such a way as to reduce the number of pathogen transmissions, such as quarantines, lockdowns, and curfews. It is very difficult to estimate a-priori the impact of such measures[32], so using a real-time indicator of social activity level can be of great help to parametrize predictive models. The correlation of the sociability parameter and road mobility resulted significant, and it was further increased by considering a very limited number of additive shifts to the sociability parameter, related to significant events related to the epidemic. Assuming that with appropriate normalization the mobility index estimates the social activity, i.e. the number of contacts, we hypothesize that the shift between mobility and sociability quantifies the gap between total contacts and unprotected contacts (i.e., potentially infectious). In fact, the shift is zero in the early period when masks and distancing were not yet widespread, and has a small positive value in summer 2021, corresponding to a likely lower attention to containment measures. Conversely, the shift is larger under stricter containment measures, i.e. the winter periods.

The second result is that we could identify the key factors affecting the number of hospitalizations along the epidemics evolution. We observed a progressive decrease in the number of hospitalizations per infection, in correspondence with the increase in vaccination coverage at first, and after the emergence of Omicron lineage secondly. In particular, after the emergence of Omicron, we observed a decoupling between the clinical records of COVID-19 epidemic and environmental measurements. While the number of infections decreased, together with the sociability index, the sewage viral load and the mobility index remained near their maxima even at the end of 2022. This result again motivates the need to complement clinical tracing with other tools to monitor social activity and the actual spread of the pathogen in the population. The decoupling between virus load and clinical outcomes is a clear sign of reduced severity, but it also shows how an epidemics can become “silent” in terms of impact on population while remaining present in the environment [33]. The need to introduce some shifts to improve the correlation between mobility data and epidemiological parameters confirms the limited ability of mobility data alone to predict long-term epidemic dynamics, as observed in [6, 7], if not complemented with other information. Anyway, the number of required shifts was very limited over an arc of 2.5 years and were likely associated to changing infection control measures.

In conclusion, our work confirms (a) the efficacy of vaccines in protecting against severe disease, and (b) that standard surveillance metrics can be inaccurate in estimating the extent of the epidemic, as already observed in [19]. In addition, (c) we show how other often available urban data, such as road traffic, allow better quantification of the pathogen transmission due to social activity, disentangling it from pathogen infectivity. The latter result also shows how mobility data can be used to predict fluctuations in social activity over short periods, and to forecast the number of new infections in support of local Public Health agencies. In a context of growing availability of open source data at urban level, that can feed models such as digital twins of a whole city[34, 35], our work shows a possible application of such models in a public health perspective, integrating social, clinical and microbiological data for enhanced surveillance of epidemic spread and evolution.

## 5 Acknowledgments

We acknowledge the Bologna MODELS4COVID Study Group of the University of Bologna and the National Institute for Nuclear Physics (INFN): Valerio Carelli, Paolo Tubertini, Luca Clissa, Stefano Diciotti,Michela Milano, Luca Palmerini, Giulia Roli, Michele Scagliarini, Rossella Miglio, Roberto Spighi, Vincenzo Vagnoni, Antonio Zoccoli, Lorenzo Chiari. We acknowledge HERA group Spa for support in sample acquisition at their sewage treatment plant for Bologna metropolitan area. We thank Prof. Frank Aarestrup and Miranda De Graaf for useful discussion about how to design the study and the sewage analysis.

## 7 Author contributions statement

E.L. implemented the mathematical model, F.D. and G.Co. analyzed the results, G.G. and V.S. performed genomic analyses, A. de C., C.C. and F.P. performed sewage sampling and analysis, G.Ca., D.R. and A.B. supported mathematical modeling and statistical data analysis and coordinated the study. All authors, including Bologna MODELS4COVID Study Group, wrote and reviewed the manuscript.

## 8 Ethical declaration

Ethical approval or informed consent were not required because the study has been performed using exclusively anonymized, leftover samples derived from the routine diagnostic procedures. The anonymization of genomic data was achieved by using the current procedure (AVR-PPC P09, rev.2) checked by the AUSL Romagna Ethical Committee (Italy).

## 9 Data availability

Data and code are hosted at https://github.com/FraDurazzi/BolognaCOVID_Sewage_Mobility.

## References

[1] Emily Howerton, Lucie Contamin, Luke C. Mullany, Michelle Qin, Nicholas G. Reich, Samantha Bents, Rebecca K. Borchering, Sung-mok Jung, Sara L. Loo, Claire P. Smith, John Levan-der, Jessica Kerr, J. Espino, Willem G. van Panhuis, Harry Hochheiser, Marta Galanti, Teresa Yamana, Sen Pei, Jeffrey Shaman, Kaitlin Rainwater-Lovett, Matt Kinsey, Kate Tallaksen, Shelby Wilson, Lauren Shin, Joseph C. Lemaitre, Joshua Kaminsky, Juan Dent Hulse, Elizabeth C. Lee, Clifton D. McKee, Alison Hill, Dean Karlen, Matteo Chinazzi, Jessica T. Davis, Kunpeng Mu, Xinyue Xiong, Ana Pastore y Piontti, Alessandro Vespignani, Erik T. Rosenstrom, Julie S. Ivy, Maria E. Mayorga, Julie L. Swann, Guido España, Sean Cavany, Sean Moore, Alex Perkins, Thomas Hladish, Alexander Pillai, Kok Ben Toh, Ira Longini, Shi Chen, Rajib Paul, Daniel Janies, Jean-Claude Thill, Anass Bouchnita, Kaiming Bi, Michael Lachmann, Spencer J. Fox, Lauren Ancel Meyers, Ajitesh Srivastava, Przemyslaw Porebski, Srini Venkatramanan, Aniruddha Adiga, Bryan Lewis, Brian Klahn, Joseph Outten, Benjamin Hurt, Jiangzhuo Chen, Henning Mortveit, Amanda Wilson, Madhav Marathe, Stefan Hoops, Parantapa Bhattacharya, Dustin Machi, Betsy L. Cadwell, Jessica M. Healy, Rachel B. Slayton, Michael A. Johansson, Matthew Biggerstaff, Shaun Truelove, Michael C. Runge, Katriona Shea, Cécile Viboud, and Justin Lessler. Evaluation of the US COVID-19 Scenario Modeling Hub for informing pandemic response under uncertainty. Nature Communications, 14(1):7260, November 2023. Number: 1 Publisher: Nature Publishing Group.

[2] Michele Tizzoni, Paolo Bajardi, Adeline Decuyper, Guillaume Kon Kam King, Christian M. Schneider, Vincent Blondel, Zbigniew Smoreda, Marta C. González, and Vittoria Colizza. On the Use of Human Mobility Proxies for Modeling Epidemics. PLOS Computational Biology, 10(7):e1003716, July 2014. Publisher: Public Library of Science.

[3] Amy Wesolowski, Caroline O. Buckee, Kenth Engø-Monsen, and C. J. E. Metcalf. Connecting Mobility to Infectious Diseases: The Promise and Limits of Mobile Phone Data. The Journal of Infectious Diseases, 214(Suppl 4):S414–S420, December 2016.

[4] Zhang Mengxi, Wang Siqin, Hu Tao, Fu Xiaokang, Wang Xioyue, Briana Halloran, Li Zhenlong, Cui Yunhe, Liu Haokun, Liu Zhimin, and Shuming Bao. Human mobility and COVID-19 transmission: a systematic review and future directions. Annals of GIS, 28(4):501–514, 2022.

[5] Serina Chang, Emma Pierson, Pang Wei Koh, Jaline Gerardin, Beth Redbird, David Grusky, and Jure Leskovec. Mobility network models of COVID-19 explain inequities and inform reopening. Nature, 2020.

[6] Federico Delussu, Michele Tizzoni, and Laetitia Gauvin. The limits of human mobility traces to predict the spread of COVID-19: A transfer entropy approach. PNAS nexus, 2(10):pgad302, October 2023.

[7] Nishant Kishore, Aimee R. Taylor, Pierre E. Jacob, Navin Vembar, Ted Cohen, Caroline O. Buckee, and Nicolas A. Menzies. Evaluating the reliability of mobility metrics from aggregated mobile phone data as proxies for SARS-CoV-2 transmission in the USA: a population-based study. The Lancet Digital Health, 4(1):e27–e36, January 2022. Publisher: Elsevier.

[8] Marino Gatto, Enrico Bertuzzo, Lorenzo Mari, Stefano Miccoli, Luca Carraro, Renato Casagrandi, and Andrea Rinaldo. Spread and dynamics of the COVID-19 epidemic in Italy: Effects of emergency containment measures. Proceedings of the National Academy of Sciences, page 202004978, April 2020. Publisher: Proceedings of the National Academy of Sciences.

[9] Matteo Chiara, Anna Maria D’Erchia, Carmela Gissi, Caterina Manzari, Antonio Parisi, Nicoletta Resta, Federico Zambelli, Ernesto Picardi, Giulio Pavesi, David S Horner, and Graziano Pesole. Next generation sequencing of SARS-CoV-2 genomes: challenges, applications and opportunities. Briefings in Bioinformatics, 22(2):616–630, March 2021.

[10] Bas B. Oude Munnink, David F. Nieuwenhuijse, Mart Stein, Áine O’Toole, Manon Haverkate, Madelief Mollers, Sandra K. Kamga, Claudia Schapendonk, Mark Pronk, Pascal Lexmond, Anne van der Linden, Theo Bestebroer, Irina Chestakova, Ronald J. Overmars, Stefan van Nieuwkoop, Richard Molenkamp, Annemiek A. van der Eijk, Corine GeurtsvanKessel, Harry Vennema, Adam Meijer, Andrew Rambaut, Jaap van Dissel, Reina S. Sikkema, Aura Timen, and Marion Koopmans. Rapid SARS-CoV-2 whole-genome sequencing and analysis for informed public health decision-making in the Netherlands. Nature Medicine, 26(9):1405–1410, September 2020. Number: 9 Publisher: Nature Publishing Group.

[11] Zhiyuan Chen, Andrew S. Azman, Xinhua Chen, Junyi Zou, Yuyang Tian, Ruijia Sun, Xiangyanyu Xu, Yani Wu, Wanying Lu, Shijia Ge, Zeyao Zhao, Juan Yang, Daniel T. Leung, Daryl B. Domman, and Hongjie Yu. Global landscape of SARS-CoV-2 genomic surveillance and data sharing. Nature Genetics, 54(4):499–507, April 2022. Number: 4 Publisher: Nature Publishing Group.

[12] The COVID-19 Genomics UK (COG-UK) consortium. An integrated national scale SARS-CoV-2 genomic surveillance network. The Lancet. Microbe, 1(3):e99–e100, July 2020.

[13] Francesco Durazzi, François Pichard, Daniel Remondini, and Marcel Salathé. Dynamics of social media behavior before and after SARS-CoV-2 infection. Frontiers in Public Health, 10, 2023.

[14] Carmela Troncoso, Dan Bogdanov, Edouard Bugnion, Sylvain Chatel, Cas Cremers, Seda Gürses, Jean-Pierre Hubaux, Dennis Jackson, James R. Larus, Wouter Lueks, Rui Oliveira, Mathias Payer, Bart Preneel, Apostolos Pyrgelis, Marcel Salathé, Theresa Stadler, and Michael Veale. Deploying decentralized, privacy-preserving proximity tracing. Communications of the ACM, 65(9):48–57, August 2022.

[15] Ray Izquierdo-Lara, Goffe Elsinga, Leo Heijnen, Bas B. Oude Munnink, Claudia M.E. Schapendonk, David Nieuwenhuijse, Matthijs Kon, Lu Lu, Frank M. Aarestrup, Samantha Lycett, Gertjan Medema, Marion P.G. Koopmans, and Miranda de Graaf. Monitoring SARS-CoV-2 Circulation and Diversity through Community Wastewater Sequencing, the Netherlands and Belgium. Emerging Infectious Diseases, 27(5):1405–1415, May 2021.

[16] Tatiana Prado, Tulio Machado Fumian, Camille Ferreira Mannarino, Paola Cristina Resende, Fernando Couto Motta, Ana Lucia Fontes Eppinghaus, Vitor Hugo Chagas do Vale, Ricardo Marinho Soares Braz, Juliana da Silva Ribeiro de Andrade, Adriana Gonçalves Maranhüo, and Marize Pereira Miagostovich. Wastewater-based epidemiology as a useful tool to track SARS-CoV-2 and support public health policies at municipal level in Brazil. Water Research, 191:116810, March 2021.

[17] Alexander Crits-Christoph, Rose S. Kantor, Matthew R. Olm, Oscar N. Whitney, Basem Al-Shayeb, Yue Clare Lou, Avi Flamholz, Lauren C. Kennedy, Hannah Greenwald, Adrian Hinkle, Jonathan Hetzel, Sara Spitzer, Jeffery Koble, Asako Tan, Fred Hyde, Gary Schroth, Scott Kuersten, Jillian F. Banfield, and Kara L. Nelson. Genome Sequencing of Sewage Detects Regionally Prevalent SARS-CoV-2 Variants. mBio, 12(1):10.1128/mbio.02703–20, January 2021. Publisher: American Society for Microbiology.

[18] Rajindra Napit, Prajwol Manandhar, Ashok Chaudhary, Bishwo Shrestha, Ajit Poudel, Roji Raut, Saman Pradhan, Samita Raut, Pragun G. Rajbhandari, Anupama Gurung, Rajesh M. Rajbhandari, Sameer M. Dixit, Jessica S. Schwind, Christine K. Johnson, Jonna K. Mazet, and Dibesh B. Karmacharya. Rapid genomic surveillance of SARS-CoV-2 in a dense urban community of Kathmandu Valley using sewage samples. PLOS ONE, 18(3):e0283664, March 2023. Publisher: Public Library of Science.

[19] Giovanni Nattino, Sara Castiglioni, Danilo Cereda, Petra Giulia Della Valle, Laura Pellegrinelli, Guido Bertolini, and Elena Pariani. Association Between SARS-CoV-2 Viral Load in Wastewater and Reported Cases, Hospitalizations, and Vaccinations in Milan, March 2020 to November 2021. JAMA, 327(19):1922–1924, May 2022.

[20] Sofia Raponi, Francesco Durazzi, Nicolas Riccardo Derus, Enrico Giampieri, Rossella Miglio, Gastone Castellani, Claudia Sala, and Bologna MODELS4COVID Study Group. Risk Factors for Admission into COVID-19 General Wards, Sub-Intensive and Intensive Care Units among SARS-CoV-2 Positive Subjects in the Municipality of Bologna, Italy, July 2023. Pages: 2023.07.12.23292559.

[21] Jessica P. Ridgway, Samuel Tideman, Bill Wright, and Ari Robicsek. Decreased Risk of Coronavirus Disease 2019-Related Hospitalization Associated With the Omicron Variant of Severe Acute Respiratory Syndrome Coronavirus 2. Open Forum Infectious Diseases, 9(7):ofac288, July 2022.

[22] Comune di Bologna. Rilevazione flusso veicoli tramite spire - anno 2021.

[23] Armando Bazzani, Enrico Lunedei, and Sandro Rambaldi. A stochastic compartmental model to simulate the Covid-19 epidemic spread on a simple network. Theoretical Biology Forum, 113(1-2):31–46, January 2020.

[24] Luca Dell’Anna. Solvable delay model for epidemic spreading: the case of Covid-19 in Italy. Scientific Reports, 10(1):15763, September 2020. Publisher: Nature Publishing Group.

[25] F. A. Rihan and H. J. Alsakaji. Dynamics of a stochastic delay differential model for COVID-19 infection with asymptomatic infected and interacting people: Case study in the UAE. Results in Physics, 28:104658, September 2021.

[26] Nicola Guglielmi, Elisa Iacomini, and Alex Viguerie. Delay differential equations for the spatially resolved simulation of epidemics with specific application to COVID-19. Mathematical Methods in the Applied Sciences, 45(8):4752–4771, May 2022.

[27] Ziren Chen, Lin Feng, Harold A. Lay, Khaled Furati, and Abdul Khaliq. SEIR model with unreported infected population and dynamic parameters for the spread of COVID-19. Mathematics and Computers in Simulation, 198:31–46, August 2022.

[28] Ruiyun Li, Sen Pei, Bin Chen, Yimeng Song, Tao Zhang, Wan Yang, and Jeffrey Shaman. Substantial undocumented infection facilitates the rapid dissemination of novel coronavirus (SARS-CoV-2). Science (New York, N.Y.), 368(6490):489–493, May 2020.

[29] James Holland Jones. Notes on R0, April 2021.

[30] Stephen A. Lauer, Kyra H. Grantz, Qifang Bi, Forrest K. Jones, Qulu Zheng, Hannah R. Meredith, Andrew S. Azman, Nicholas G. Reich, and Justin Lessler. The Incubation Period of Coronavirus Disease 2019 (COVID-19) From Publicly Reported Confirmed Cases: Estimation and Application. Annals of Internal Medicine, 172(9):577–582, May 2020.

[31] Md. Arif Billah, Md. Mamun Miah, and Md. Nuruzzaman Khan. Reproductive number of coronavirus: A systematic review and meta-analysis based on global level evidence. PLoS ONE, 15(11):e0242128, November 2020.

[32] Roy M. Anderson, Hans Heesterbeek, Don Klinkenberg, and T Déirdre Hollingsworth. How will country-based mitigation measures influence the course of the COVID-19 epidemic? The Lancet, 395(10228):931–934, 2020.

[33] Frank M. Aarestrup, Marc Bonten, and Marion Koopmans. Pandemics– One Health preparedness for the next. The Lancet Regional Health – Europe, 9, October 2021. Publisher: Elsevier.

[34] Daoguang Yang, Hamid Reza Karimi, Okyay Kaynak, and Shen Yin. Developments of digital twin technologies in industrial, smart city and healthcare sectors: a survey. Complex Engineering Systems, 2021.

[35] Nils Chr Stenseth, Rudolf Schlatte, Xiaoli Liu, Roger Pielke, Ruiyun Li, Bin Chen, Ottar N. Bjørnstad, Dimitri Kusnezov, George F. Gao, Christophe Fraser, Jason D. Whittington, Yuqi Bai, Ke Deng, Peng Gong, Dabo Guan, Yixiong Xiao, Bing Xu, and Einar Broch Johnsen. How to avoid a local epidemic becoming a global pandemic. Proceedings of the National Academy of Sciences of the United States of America, 120(10):e2220080120, March 2023.

