## Supplementary Information for "Human mobility and sewage data correlate with COVID-19 epidemic evolution in the metropolitan area of Bologna"

† Members list can be found in the Acknowledgments section.

### Supplementary Text

**Basic reproduction number $R_0$ and estimation of the associated parameters**

Following [1], $R_0$ is defined as the expected number of secondary cases produced by a single infection in a completely susceptible population. For this reason, it can be estimated with rigor only at the beginning of an outbreak. More specifically $R_0 = i \cdot c \cdot T$ , where *i* is the infectivity (i.e. probability of infection given a contact between a susceptible and an infected in individual), *c* is the average rate of contacts between susceptible and infected individuals and *T* is the duration of the infectiousness ($T_U$ in our model). In our model, the β parameter we define stands for the term $i \cdot c$ shown here, describing together both transmissivity and social contacts, thus we refer to it as *rate of potentially infectious contacts*. To model separately the variation of infectivity and sociability over time, we introduce two multiplicative coefficients as in $\beta = s \cdot \tau \cdot \beta_0$, where τ stands for the relative infectivity w.r.t. the initial SARS-CoV-2 variant, accounting for virus evolution over time, and *s* stands for the relative sociability, which describes the variation of the average number of social contacts with respect to its initial value. In the Methods section of the paper we explain how τ has been modified over time, according to the information about infectivity of the prevalent SARS-CoV-2 variants.

The sociability parameter *s* was estimated with weekly frequency by 1) defining an appropriate interval of *s* values to start from, based on value of s at previous 1-week time window; 2) calculating the evolution of the epidemiologic model as a function of the defined range of values for the sociability parameter *s*, and 3) identifying the value of *s* minimizing the square error between the observed number of new cases and the model output over the last week of available measurements (corresponding to 7 daily time points). For all estimations, a step of 1 × 10^-2 has been used to span the initial s range, thus all our *s* estimations have the same uncertainty of ±0.01 which represents the resolution limit of the parameter estimate. Considering the full duration of the study presented in the paper (866 time points), the MAE on estimated cases is 42 (RMSE = 97, see Supplementary Fig. S1).

**Description of COVID-19 infections in the compartmental model**

Our model assumes 7.5 days ($T_E$+$T_U$) from infection to isolation, the first 2 being in the Exposed compartment ($T_E$=2) and the remaining 5.5 in the Unreported infected compartment ($T_U$=5.5). During the first 2 days in the Exposed compartment, individuals are not infectious. Then, during their permanence in the Unreported infected compartment, they can infect Susceptible individuals. Since we are not modelling clinical outcomes, we are not explicitly modelling an incubation period, defined as time from infection to symptoms development. Anyway, this period with a median duration of 5.5 days[2] corresponds in our model to the first 2 not-infectious days ($T_E$) plus 3.5 days as an infectious Unreported infected individual. After 2 additional days in the U compartment, with which we model delays in symptoms acknowledgment and testing, an Unreported infected individual may become Isolated.

**Change point detection of the relationship between sociability parameter and mobility index**

In the main document, we identified 3 time points to re-align the sociability parameter and the mobility index by adding a constant term to its time series. The choice of the exact date was guided by the knowledge of significant external events that could have changed the relationship between human mobility (described by the mobility index *m*) and the effective number of infectious contacts (described by the sociability parameter *s*). Here, we show the results of a data-driven change point detection approach. We first computed the time series of the difference between the sociability parameter s and the mobility index m at weekly intervals, and then operated change point detection with *ruptures* Python package [3], using the mean squared difference between time points and the average in each segment as a minimization objective, and requiring a minimum of 13 weeks (3 months) for each segment. The 4 change points identified by this analysis are listed in Supplementary Table S2. Each time point is less than 30 days from the manually curated breakpoints selected in the main document (2 over 3 time points are less that 2 weeks distant in time). The additional breakpoint is set on 21$^{st}$ March 2022, in close correspondence with Omicron lineage emergence (19$^{th}$ March), which we already commented as a the time point after which the correlation between sociability and mobility was getting lower, also related to the loss of correlation between viral load in sewage and clinical cases.

This analysis shows that It could be in principle possible to identify the main shifts between sociability parameter and mobility index also through statistical analysis, but since we have studied only one example of this approach with very few breakpoints (i.e. the COVID-19 epidemic with 3 break points identified in about two years of follow-up), we do not claim that these results are sufficiently validated to be used in every future context without additional knowledge (e.g. about the most relevant interventions on social restrictions).

**Modelling reduced vaccine efficacy**

In the model presented in the main text, we implement vaccination with 100% efficacy (complete immunization of the vaccinated individuals). Since this assumption may not hold, we have rerun the simulations considering a 75% efficacy for the vaccine (thus 25% vaccines have no effects). These new simulations (see Supplementary Fig. S4) show that 1) there are some differences only in a limited temporal region, namely around the infections peak in summer 2021, while since October 2021 the outputs (i.e. the estimated sociability parameter *s)* overlap with the previous results; 2) the match between sociability and mobility parameters requires a change in the value of the additive shift from +0.23 to +0.43 only in the period from 17$^{th}$ May to 15$^{th}$ September 2021 (in the previous period vaccines were not available), while the additive shift remains substantially the same in the following period (from +0.71 to +0.74). Thus, even in a model with reduced vaccine efficacy, we can observe that the trends of mobility and sociability parameters remain highly correlated up to an additive constant, that has to be added only in correspondence to specific events like lockdown measures.

### Supplementary Tables

| | Phase A | Phase B | Phase C |
|---|---|---|---|
| $r_{CV}$ | 0.5 ± 0.3 | 0.5 ± 0.2 | 0.19 ± 0.19 |
| $r_{HC}$ | 730 ± 30 | 257 ± 15 | 36 ± 3 |

**Table S1.** MCMC estimates of the parameters shown in Figure 3, with standard deviation of the MCMC trace as error.

| Manually curated breakpoints | Statistically detected change-points |
|---|---|
| 18/05/2020 | 05/05/2020 |
| 17/05/2021 | 19/06/2020 |
| 15/09/2021 | 27/09/2021 |
| | 21/03/2022 |

**Table S2.** Left column: breakpoints used for the analysis in the main document. Right column: change points detected as detailed in the Supplementary Text.

### Supplementary Figures

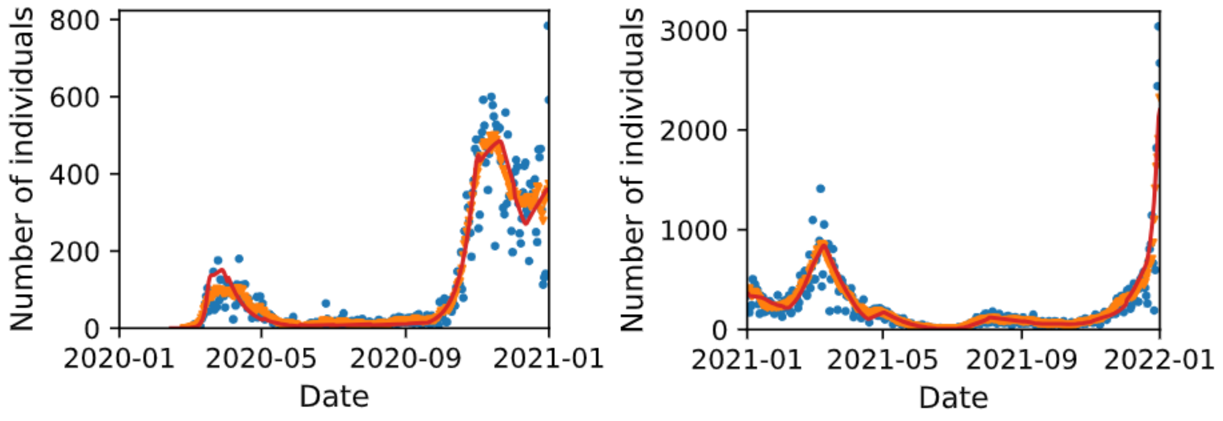


(a) (b)

**Figure S1.1.** Comparison of clinical data (blue dots = number of cases, orange dots = moving average) and model output (red line), fitted to the data through the sociability parameter *s*. (a) year 2020, (b) year 2021, separated for visibility because of different scales on the y axis.

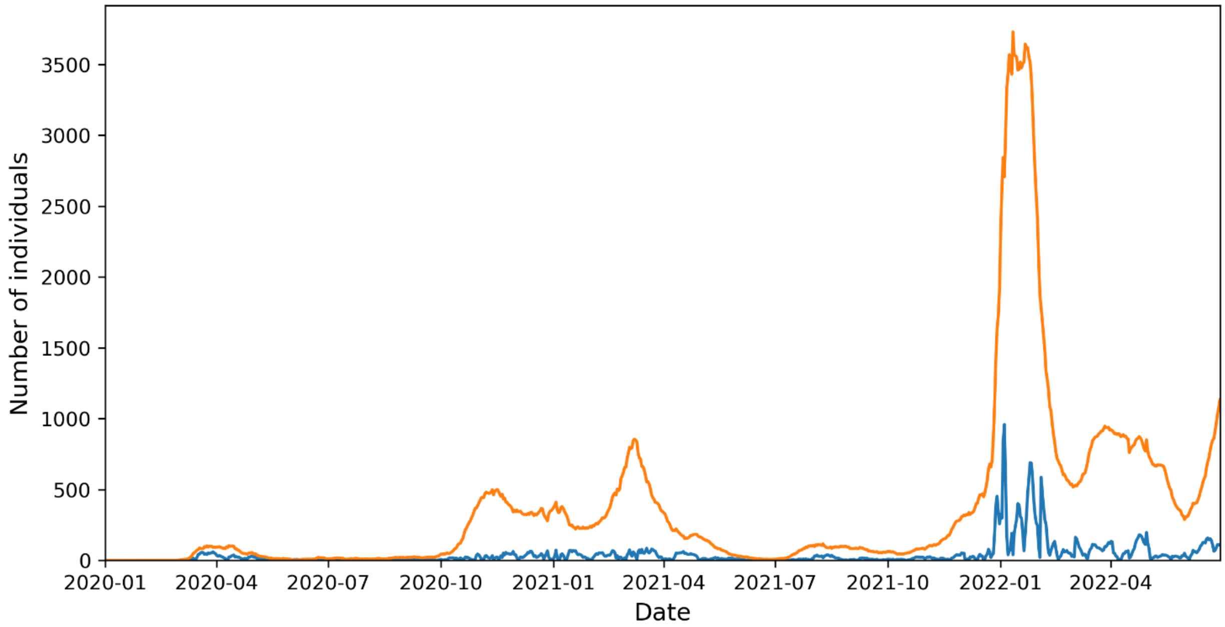


**Figure S1.2.** Time series of the weekly moving average of observed new cases (orange line) and of the Absolute Error (blue line) between the predicted new cases and the moving average of the observed ones. The Mean Absolute Error (MAE) achieved by the model is 42 (RMSE = 97) over a study duration of 866 days.

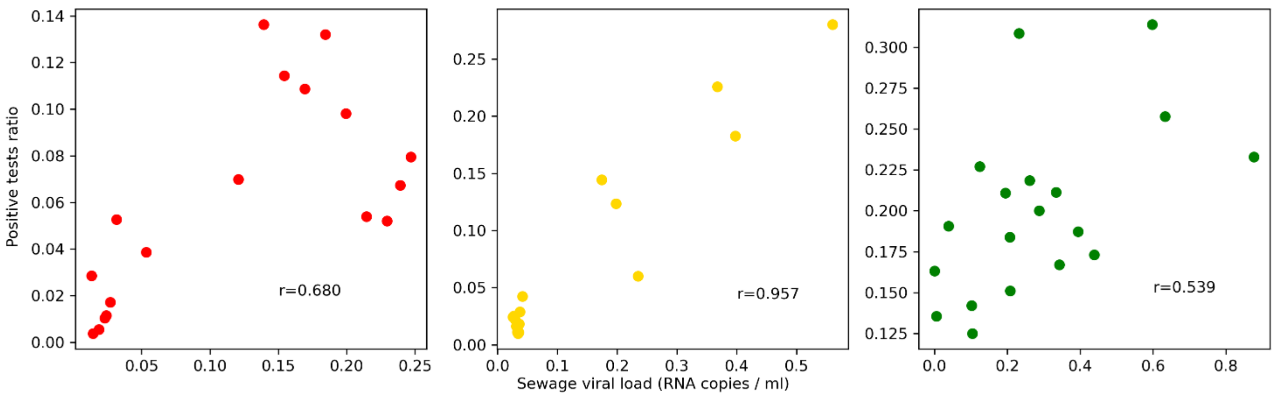


(a)

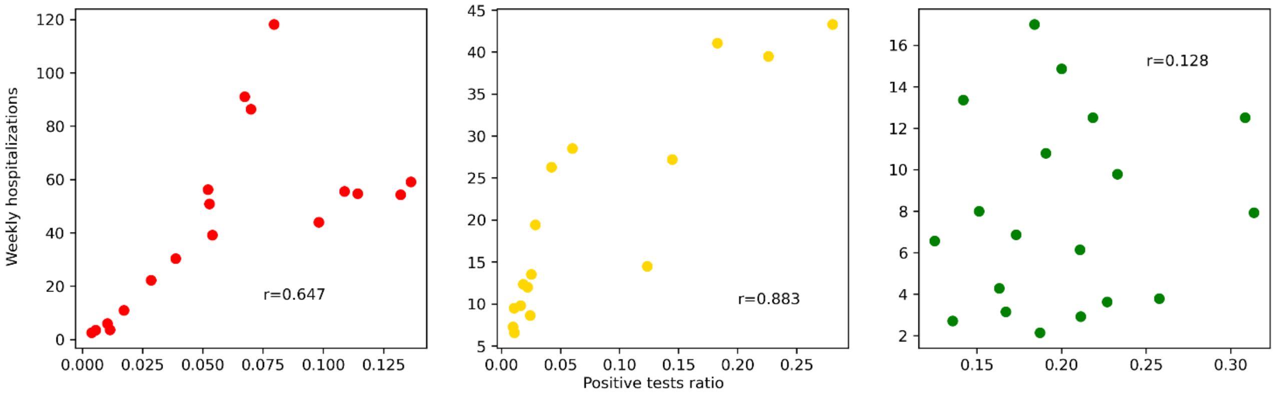


(b)

**Figure S2.** Scatter plot of (a, top) the sewage viral load vs positive test ratio and (b, bottom) positive test ratio vs weekly hospitalizations. Pearson's linear correlation coefficient r is reported separately for each time phase in which we divided our time series. We tested the Pearson's linear correlation between the two pairs of time series with a time lag of -1, 0, 1, 2 weeks, and the maximum values were achieved with a 0-week lag (data not shown).

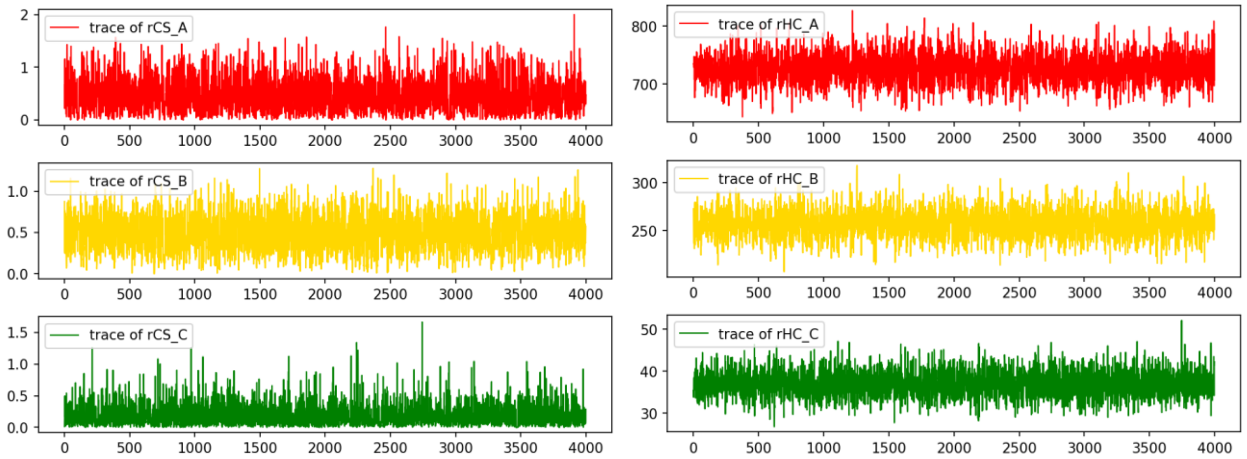


**Figure S3.** Trace of the parameters sampled with Monte Carlo Markov chain from pymc3. Each parameter has been sampled with 4000 steps (4 chains of 1000 steps, shown concatenated in each subplot). The sample acceptance probability (*target_accept* parameter in pymc3 package) has been set to 0.85: it yielded 0 observed divergences in the sampling, confirming the stability of the algorithm convergence. We observe how convergence of MCMC model is also motivated by the absence of drift and by the similarity of the 4 concatenated 1000-step traces.

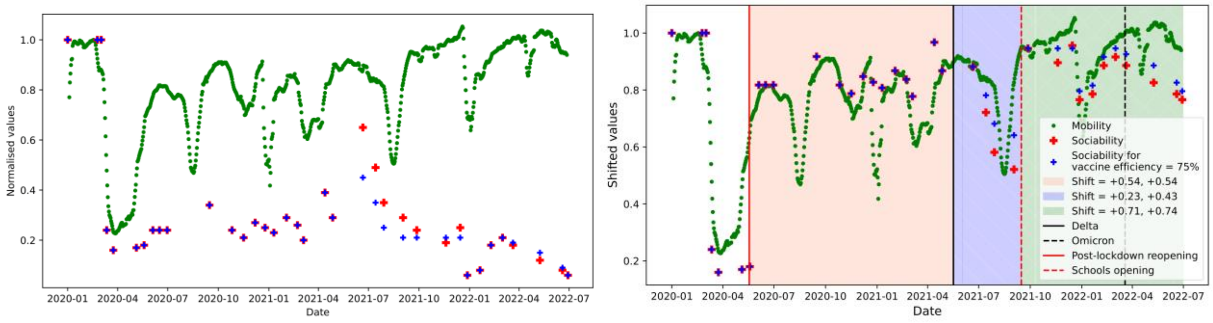


(a) (b)

**Figure S4.** Estimates of sociability *s* parameter with (red) 100% vaccine efficacy and (blue) 75% efficacy. a) Normalized mobility (green) and sociability over time. b) Normalized mobility and shifted sociability values as in Figure 4.

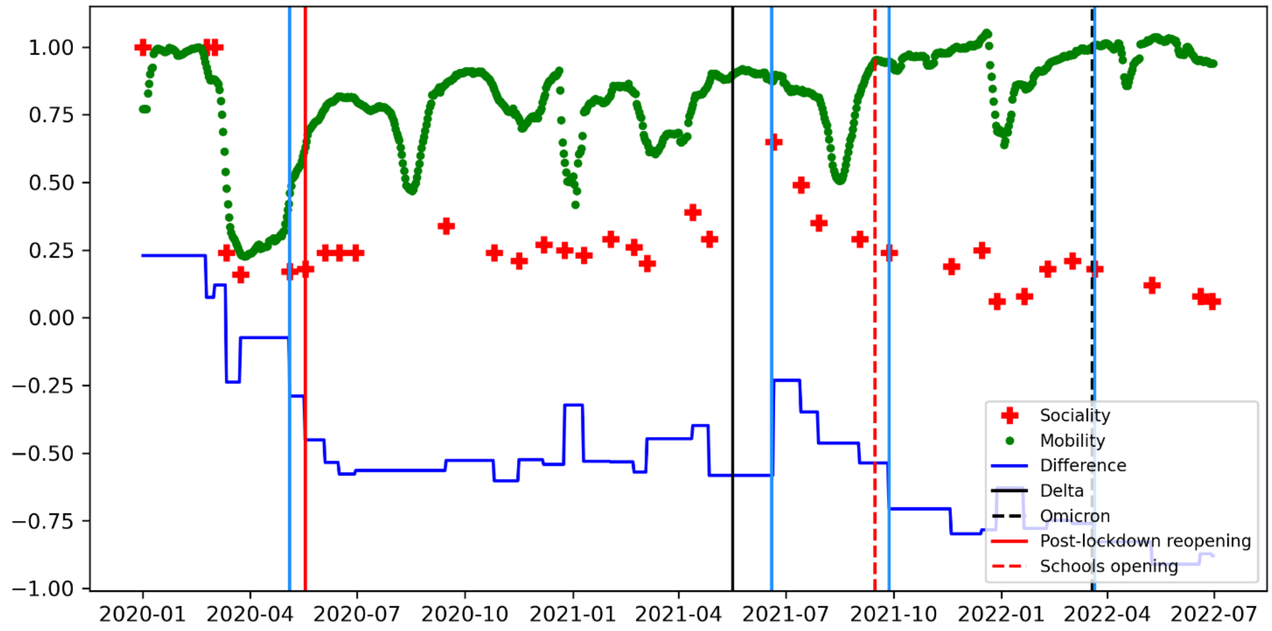


**Figure S3.** *Sociability parameter vs Mobility index*. The blue line represents the difference between the sociability parameter *s* (red crosses) and the mobility index *m* (green dots). The change points identified by statistical breakpoint analysis are represented as light blue vertical lines. The remaining vertical lines correspond to manually selected breakpoints guided by external relevant events (different colors corresponding to different types of events, see figure legend). We remark that the last breakpoint corresponding to Omicron arrival was not considered.

### References

[1] J. Holland Jones, «Notes on R0». 5 aprile 2021. [Online]. Disponibile su: https://populationsciences.berkeley.edu/wp-content/uploads/2021/06/Jones-Notes-on-R0.pdf

[2] S. A. Lauer *et al.*, «The Incubation Period of Coronavirus Disease 2019 (COVID-19) From Publicly Reported Confirmed Cases: Estimation and Application», *Ann. Intern. Med.*, vol. 172, fasc. 9, pp. 577–582, mag. 2020, doi: 10.7326/M20-0504.

[3] C. Truong, L. Oudre, e N. Vayatis, «Selective review of offline change point detection methods», *Signal Process.*, vol. 167, p. 107299, feb. 2020, doi: 10.1016/j.sigpro.2019.107299.
